# The impact of the COVID-19 pandemic on college students’ health and financial stability in New York City: Findings from a population-based sample of City University of New York (CUNY) students

**DOI:** 10.1101/2020.12.08.20246074

**Authors:** Heidi E. Jones, Meredith Manze, Victoria Ngo, Patricia Lamberson, Nicholas Freudenberg

## Abstract

Understanding the effect of the coronavirus disease 2019 (COVID-19) pandemic on students’ health and financial stability is important to establish effective interventions to mitigate these effects, which may have long-term consequences on their health and wellbeing. Public universities in urban centers represent a substantial proportion of college students in the United States. We implemented a cross-sectional population-based online survey of 2,282 students in a large, public university in New York City in April 2020. We created weights to account for non-response and used Poisson regression with robust standard errors to estimate adjusted prevalence ratios (aPR) for factors associated with mental health outcomes. Students experienced high rates of anxiety/depression and financial instability due to the pandemic. Half of the students reported anxiety/depression (54.5%) and an increased need for mental health services (49.0%) as a result of the COVID-19 pandemic. The majority (81.1%) reported loss of household income, and half (49.8%) reported worries about losing housing. High levels of food (aPR=1.4, 95% CI 1.2, 1.6) and housing (aPR=1.3, 95% CI 1.2, 1.4) insecurity were the strongest predictors of anxiety/depression. Household and personal experiences with possible COVID-19 symptoms were also associated with anxiety/depression or the need for increased mental health services. Addressing student needs at public urban universities requires an integrated holistic approach that targets urgent mental health and economic needs related to the impact of COVID-19. Students who become infected need mental health services as well as health monitoring.

## Introduction

In the United States (US), New York City (NYC) became the first epicenter of the coronavirus disease 2019 (COVID-19) pandemic in late March 2020.^1^ During this first wave in NYC, the epidemic peaked with an estimated seven-day rolling average high of over 5,200 cases in early April 2020 and a death rate of 228 confirmed COVID-19 deaths per 100,000 people between March and August 2020;^2^ the true death rate is likely higher.^3^ The primary public health response to the pandemic promoted social distancing to reduce community transmission, including shifting to online learning environments for schools and college campuses. The City University of New York (CUNY), a public university with 25 campuses across the five boroughs of NYC, officially closed its physical facilities and began virtual classes on March 19, 2020 in response to the pandemic.^4^

In 2019-2020, CUNY enrolled a large, diverse student body, with an estimated 274,000 matriculated students in programs that offer associate, baccalaureate or graduate degrees. Many CUNY students come from low income households; in 2016, an estimated three in five CUNY undergraduates lived in households with annual incomes of less than $30,000.^5^ At the same time, a number of CUNY campuses rank highly among US colleges for the upward financial mobility of their graduates.^6^ Further, about 40% of New York City’s 689,000 college students are enrolled at CUNY.^7^ Understanding how the COVID-19 pandemic has affected their health and wellbeing can inform interventions and policies to mitigate harm, with implications for health, equity, economic development, higher education and social justice in the region. These findings may also be applicable to other large public urban universities across the country. We sought to investigate the impact of the COVID-19 pandemic on CUNY students’ self-reported general and mental health, as well as on their financial stability at the height of the first wave of the epidemic in NYC.

## Methods

We conducted a cross-sectional online survey of a representative sample of CUNY students from April 14-22, 2020. The study was reviewed and approved by the CUNY School of Public Health’s Institutional Review Board (Protocol #695980); the survey began with informed consent, requiring participants to indicate consent electronically prior to starting the survey. The first 1,000 respondents received a $20 gift card for their participation by email, and five respondents among all who participated were randomly selected to receive $100 gift cards.

### Sample

In collaboration with the CUNY Office of Institutional Research, which maintains a centralized database of CUNY students, we used simple random selection to select 10,000 students from the approximately 274,000 matriculated students in the spring 2020 semester. Of those, 2,282 completed 70% or more of the entire survey (response rate of 23%) and are included in this analysis. Given response bias, we created sampling weights to account for non-response based on sex, race/ethnicity, type of campus (undergraduate versus graduate), and full-time or part-time student status; all measures available in the CUNY central database.

### Survey instrument

The survey was pilot tested with five students to ensure comprehension and readability and was designed to take 15 minutes to complete. The survey comprised five domains: educational experience, household/living situation, economic impact, health impact and socio-demographic characteristics; other socio-demographic and student status data such as sex (categorized as male or female), race/ethnicity, campus and full-time or part-time student status were taken from the centralized student database. Items to assess health included a question on general health as recalled at the beginning of the semester and at the time of the survey taken from the Health Related Quality of Life Healthy Days Core Module (CDC HRQOL– 4),^8^ the Patient Health questionnaire-4 (PHQ-4) to measure anxiety and depression,^9, 10^ and changes in patterns of substance use. Experiences with COVID-19 symptoms and testing were also assessed. To measure economic impact, we included questions on loss of income, changes in household expenses, food insecurity (adapted from the 6-item USDA Food Security Survey),^11^ and concerns about housing costs. Each domain primarily consisted of closed ended questions; all but the socio-demographic section ended with an open-ended question for students to describe anything that was missed about their experiences in their own words. Similarly questions that allowed students to select more than one option included an ‘other’ category with space for text, in case important response categories were missed. The current analysis focuses on the health and economic impact of the COVID-19 pandemic on CUNY students; future analyses will focus on the effect on educational experiences.

### Analysis

We present weighted estimates of socio-demographic characteristics by level of students (graduate, bachelors or associate degree) as an overview of the CUNY student population. Next we present weighted estimates of students’ reports on the impact of the COVID-19 on health and economic measures. We used chi-squared tests to compare reports on changes in substance use and anticipated graduation by anxiety/depression. Finally, we conducted weighted multivariable Poisson regression models with robust standard errors to estimate the prevalence ratios for factors hypothesized to be associated with anxiety/depression, and an increased need for mental health services as a result of the pandemic. For these models, we explored bivariate associations with exposures to stressors including: concerns about losing housing, reduced ability to buy needed food, students or household members experiencing COVID-19-like symptoms, having a household member who is a healthcare worker, being a parent or guardian, being a caretaker of adult(s) over age 65, and whether students expected to graduate later than previously anticipated, as hypothesized contributors to anxiety/depression. Changes to substance use were not included, as we hypothesized that anxiety/depression may have led to increased substance use. We also explored bivariate associations with socio-demographic characteristics (sex, race/ethnicity, age) and student characteristics (associates, bachelors or graduate; full-time versus part-time student; New York City, New York state, other state or international residency status) as potential confounders of our hypothesized exposures. No independent variables in the final adjusted models were strongly correlated with one another (correlation of 0.50 or greater). Final models include exposures associated with the outcome at the bivariate level, which remained significant (p<0.05) in multivariable models, after adjusting for race/ethnicity and age. Type of degree (associates, bachelors or graduate), full-time versus part time student, residency status, being a parent/guardian, and having a family member work in healthcare were not significantly associated with anxiety/depression or increased need for mental health in fully adjusted models and were not included in the final models.

For open-ended questions or ‘other’ response categories, we identified common themes and had two independent reviewers code each quote in a blinded fashion with discussion to resolve discrepancies. We present the frequency with which themes about mental health and financial stability arose, and include illustrative quotes to contextualize the quantitative findings and illuminate the lived experiences of students.

## Results

CUNY students are generally young (66.0% 18-25 years of age) and more female than male (57.9% versus 42.1% respectively, Table 1). Over one quarter of students are Hispanic (29.4%), followed by Black non-Hispanic (24.9%), White non-Hispanic (23.0%), and Asian Pacific Islander (22.1%), with 0.4% American Indian/Native Alaskan. Students enrolled in graduate, bachelor and associate degree programs differ by socio-demographic characteristics such as female-male ratio, age and race/ethnicity (Table 1). Only 1.9% of students reported living in school dormitories prior to the pandemic, with the remainder in different types of off-campus living situations; living with a parent or relative (56.2%) or in their own house/rental apartment (34.7%) were the most common.

**Table 1:**
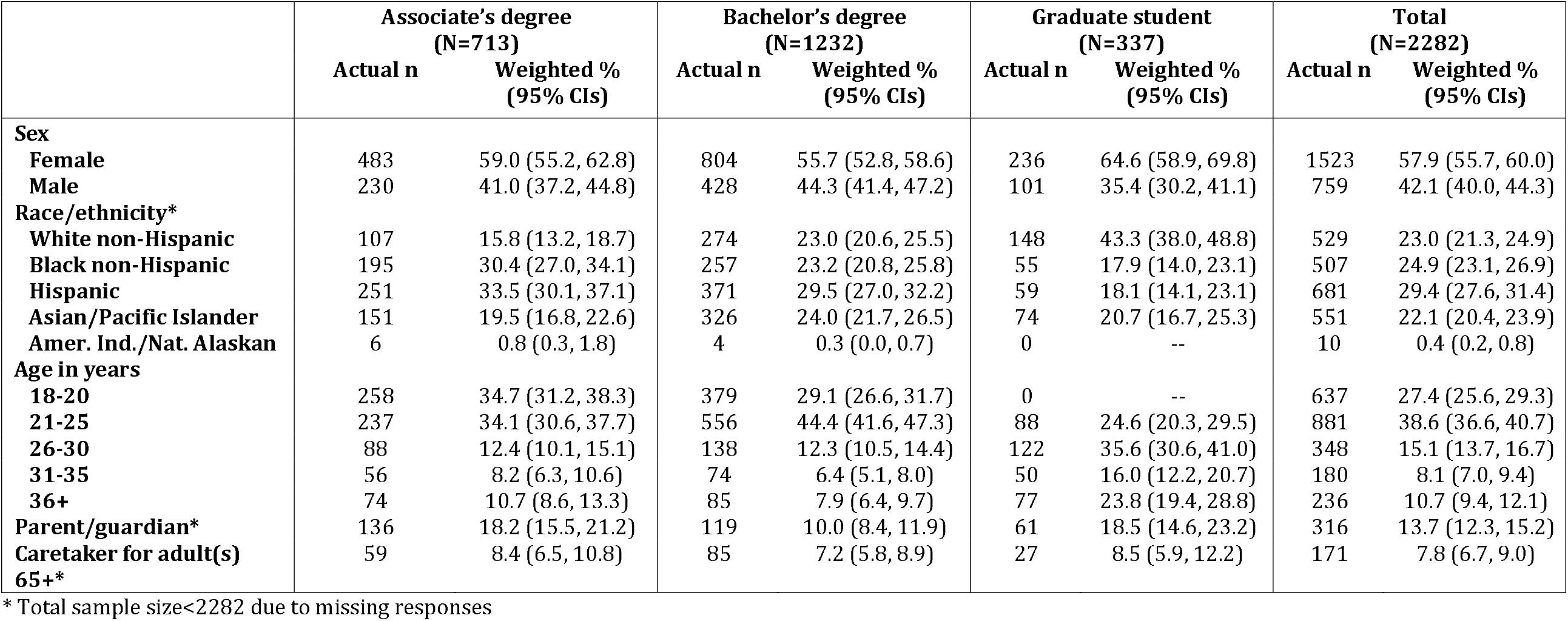
Socio-demographic characteristics by associate’s, bachelor’s or graduate student, City University of New York (CUNY) Student COVID-19 Survey, April 2020

### Health outcomes

While only 8.2% reported that their general health had been fair/poor at the beginning of the semester, 26.6% reported their general health as fair/poor at the time of the survey, (Table 2), with no differences by type of degree (not shown). More than one quarter (29.4%) reported experiencing symptoms that could be COVID-19, of whom only 15.4% were tested. Among all students, 5.8% were tested for COVID-19 of whom 29.1% reported a positive result (at the time of the survey, testing was not widely available in New York City^12^). More than a quarter (29.5%) of students also reported someone in their household had experienced symptoms that could be COVID-19.

**Table 2:** Health and financial impact of COVID-19 pandemic, City University of New York (CUNY) Student COVID-19 Survey, April 2020

|  | **Total**<br>**(N=2282)** |  |
| --- | --- | --- |
|  | **Actual n** | **Weighted %**<br>**(95% CIs)** |
| **Health and financial impact indicators** |  |  |
| **General health at beginning of semester\*** |  |  |
| Excellent/Very Good | 1038 | 68.4 (66.4, 70.3) |
| Good | 606 | 23.4 (21.7, 25.2) |
| Fair/Poor | 628 | 8.2 (7.1, 9.4) |
| **General health since COVID-19 epidemic\*** |  |  |
| Excellent/Very Good | 464 | 47.1 (45.0, 49.2) |
| Good | 606 | 26.3 (24.5, 28.2) |
| Fair/Poor | 502 | 26.6 (24.8, 28.5) |
| **Experienced COVID-19-like symptoms\*** | 663 | 29.4 (27.5, 31.4) |
| **Household member experienced COVID-19 like symptoms\*** | 658 | 29.5 (27.6, 31.5) |
| **Depression\*** | 963 | 43.2 (41.1, 45.3) |
| **Anxiety\*** | 993 | 42.2 (40.1, 44.4) |
| **Increased need for help with stress/anxiety\*** | 1116 | 49.0 (46.9, 51.2) |
| **Loss of household income\*** | 1799 | 81.1 (79.3, 82.7) |
| **Reduced household ability to buy food\*** |  |  |
| A lot | 547 | 24.1 (22.3, 26.0) |
| Somewhat | 823 | 35.9 (33.9, 38.0) |
| A little | 558 | 24.7 (22.9, 26.6) |
| Not at all/made it easier | 340 | 15.2 (13.7, 16.8) |
| **Often/sometimes worried about running out of food before able to afford more\*** | 1150 | 50.1 (48.0, 52.2) |
| **Often/sometimes skip a meal because cannot afford food\*** | 597 | 26.6 (24.8, 28.6) |
| **Often/sometimes unable to eat nutritious meals for lack of money\*** | 796 | 35.4 (33.4, 37.5) |
| **Often/sometimes have gone hungry for lack of access to food\*** | 403 | 17.9 (16.3, 19.6) |
| **How worried about losing housing\*** |  |  |
| Very worried | 317 | 14.1 (12.7, 15.7) |
| Somewhat worried | 812 | 35.7 (33.7, 37.7) |
| Not worried at all | 1143 | 50.2 (48.1, 52.3) |
| **Expect to graduate\*** |  |  |
| Later than expected | 586 | 26.2 (24.4, 28.2) |
| Don't know | 665 | 29.3 (27.4, 31.3) |
| Earlier/same time as expected | 1030 | 44.4 (42.3, 46.5) |

\* Sample size varies from 2215 to 2281 due to missing values

More than half of students (54.5%) reported experiencing anxiety and/or depression, with 43.2% reporting anxiety and 42.2% depression. Feeling overwhelmed, anxious, and/or depressed was also a common theme identified in open-ended questions. When asked, to describe other effects of the coronavirus on their educational experience in an open-ended question, 37.3% of the participants responded. Mental health was identified as a theme in 26.8% of these responses. One student described it this way, “I have this feeling of impending doom like world war three will start, I’m scared.”

Almost half (49.0%) reported an increased need for mental health services to help cope with stress, anxiety or depression due to the pandemic. Some students reported increased substance use in response to the pandemic: 13.6% reported increased usage of alcohol, 8.1% marijuana, 4.0% vaping, and 3.4% cigarette smoking. Students who had anxiety/depression were two to three times as likely to report an increase in substance use as those not reporting anxiety/depression: alcohol usage (17.8% versus 8.3%, p<0.01), marijuana use (10.6% versus 4.9%, p<.01), vaping (5.8% versus 1.9%, p<.01) and cigarette smoking (4.5% versus 2.1%, p<0.01).

More than half of students (56.8%) reported their ability to do schoolwork decreased as a result of the pandemic, of whom 73.4% identified their mental state as a reason for this decrease. This belief was also identified in open-ended responses. When asked, “What has affected your ability to do schoolwork,” 4.5% (n=102) selected “other” and provided text responses, of which 30.4% were coded as mental health-related. One student wrote, “My mind wanders too much because I am anxious.” Another student explained, “It is mentally exhausting not knowing what’s happening. Not knowing what the future holds.” One quarter (26.2%) reported expecting to graduate later than anticipated as a result of the pandemic, and another 29.3% were uncertain of its impact on graduation date (Table 2). Those experiencing anxiety/depression were more likely to report anticipating graduating later than expected than those not reporting anxiety/depression (32.1% versus 19.2%, p<0.01).

### Economic outcomes

The majority of students (81.1%) reported that they (54.1%) and/or someone else in their household (68.9%) lost income as a result of the pandemic. Nearly half (47.8%) reported their weekly household expenses increased, with 21.2% reporting a decrease in weekly expenses and 30.9% no change. The biggest increases in household expenses reported were an increase in the cost of cleaning supplies (72.9%) and food costs (71.4%), followed by an increase in the cost of support for family members outside of the household (37.4%), entertainment (30.8%), and medical costs (28.8%).

Half of the students (49.8%) reported being very or somewhat worried about losing their housing as a result of the pandemic and about half reported experiencing some forms of food insecurity in the two weeks preceding the survey (Table 2). For example, 13.2% reported often and 37.0% sometimes being worried they would run out of food before being able to afford more, and 4.5% often and 13.3% sometimes had gone hungry for lack of access to food. Sixty percent (60.0%) reported their household’s ability to purchase food had been reduced a lot (24.1%) or somewhat (35.9%) as a result of the pandemic. Food insecurity was identified as a theme in open-ended responses as well. For example, when asked about other ways the coronavirus has affected their current living situation, of the 21.6% who responded (n=493), 9.7% of the responses were coded as relating to food insecurity. One student reported, “Since the epidemic began, unfortunately only my uncle is working and we are trying to make ends meet with the rent and the utility bills. It’s been stressful because some days we eat less just to be cautious of how much money is being spent.”

Students also identified general financial concerns. Among the 21.6% who described other ways the coronavirus has affected their current living conditions, 61.1% of the responses were coded as financial concerns. One student explained, “My mom has been reduced to working 10-15 hours, risking her health to pay her rent which hasn’t been postponed. She has no health care, so my income is going to her for her to pay for food and rent.” Another student reported, “I’m not sure how long I’ll be employed. While I work additional hours online now, there is no guarantee that I’ll be working much longer. My landlady isn’t exactly understanding so there is a chance that eviction could be an issue.”

### Factors associated with anxiety/depression and increased need for mental health services

Females were 1.4 (95% confidence intervals CIs, 1.2, 1.5) times as likely as males to report anxiety/depression, with no significant differences by age (Table 3). Financial stressors—the ability to buy food reduced by a lot (aPR=1.4, 95% CIs 1.2, 1.6) and being very worried about losing housing (aPR=1.3, 95% CIs 1.2, 1.4)—and expecting to graduate later than originally planned (aPR=1.3, 95% CIs 1.1, 1.4) had the strongest associations with anxiety/depression in adjusted models. Being a caretaker for an adult over the age of 65 (aPR=1.2, 95% CIs 1.1, 1.4) and having a household member who experienced COVID-like symptoms (aPR=1.2, 95% CIs 1.1, 1.3) were also associated with anxiety/depression. Similarly, financial stressors and being female were the exposures with the strongest association with reporting an increased need for mental health support (Table 2). Reports of needing an increased level of mental health support were associated with the student experiencing COVID-like symptoms (aPR=1.2, 95% CIs 1.1, 1.3), rather than a household member experiencing symptoms.

**Table 3:** Factors associated with anxiety/depression and an increased need for mental health services, City University of New York (CUNY) Student COVID-19 Survey, April 2020

| Weighted estimates | Anxiety/Depression<br>(N=2206) |  |  | Increased need for mental health<br>services (N=2185) |  |  |
| --- | --- | --- | --- | --- | --- | --- |
|  | Weighted<br>% | Crude PR<br>(95% CI) | Adjusted PR<br>(95% CI) | Weighted<br>% | Crude PR<br>(95% CI) | Adjusted PR<br>(95% CI) |
| **Sex** |  |  |  |  |  |  |
| Female | 61.4 | 1.4 (1.2, 1.5) | 1.4 (1.2, 1.5) | 56.3 | 1.4 (1.3, 1.6) | 1.4 (1.2, 1.5) |
| Male | 45.1 | 1.0 | 1.0 | 39.1 | 1.0 | 1.0 |
| **Race/ethnicity** |  |  |  |  |  |  |
| White non-Hispanic | 56.9 | 1.0 | 1.0 | 52.4 | 1.0 | 1.0 |
| Black non-Hispanic | 49.5 | 0.9 (0.8, 1.0) | 0.8 (0.7, 0.9) | 44.0 | 0.8 (0.7, 1.0) | 1.4 (0.5, 3.8) |
| Hispanic | 58.2 | 1.0 (0.9, 1.1) | 1.1 (0.6, 1.9) | 53.2 | 1.0 (0.9, 1.1) | 1.6 (0.6, 4.6) |
| Asian/Pacific Islander | 53.0 | 0.9 (0.8, 1.0) | 1.0 (0.6, 1.8) | 46.1 | 0.9 (0.8, 1.0) | 1.4 (0.5, 4.0) |
| American Indian/Native Alaskan | 46.6 | 0.8 (0.4, 1.6) | -- | 25.8 | 0.5 (0.2, 1.4) | -- |
| **Age in years** |  |  |  |  |  |  |
| 18-20 | 51.9 | 1.0 | 1.0 | 43.6 | 1.0 | 1.0 |
| 21-25 | 55.1 | 1.1 (1.0, 1.2) | 1.0 (0.9, 1.1) | 48.9 | 1.1 (1.0, 1.3) | 1.1 (0.9, 1.2) |
| 26-30 | 57.9 | 1.1 (1.0, 1.3) | 1.0 (0.9, 1.2) | 53.2 | 1.2 (1.1, 1.4) | 1.1 (1.0, 1.3) |
| 31-35 | 57.6 | 1.1 (0.9, 1.3) | 1.0 (0.9, 1.2) | 54.7 | 1.3 (1.1, 1.5) | 1.1 (1.0, 1.3) |
| 36+ | 51.9 | 1.0 (0.9, 1.2) | 0.9 (0.8, 1.0) | 53.8 | 1.2 (1.0, 1.4) | 1.1 (1.0, 1.3) |
| **Caretaker for adult(s) aged 65+** |  |  |  |  |  |  |
| Yes | 74.2 | 1.4 (1.3, 1.6) | 1.2 (1.1, 1.4) | 59.3 | 1.2 (1.1, 1.4) | 1.0 (0.9, 1.2) |
| No | 52.9 | 1.0 | 1.0 | 48.3 | 1.0 | 1.0 |
| **Student had COVID-19-like symptoms** |  |  |  |  |  |  |
| Yes | 64.0 | 1.3 (1.2, 1.4) | 1.1 (1.0, 1.2) | 59.4 | 1.3 (1.2, 1.4) | 1.2 (1.1, 1.3) |
| No | 50.6 | 1.0 | 1.0 | 44.9 | 1.0 | 1.0 |
| **Household member had COVID-19-like symptoms** |  |  |  |  |  |  |
| Yes | 64.4 | 1.3 (1.2, 1.4) | 1.2 (1.1, 1.3) | 55.8 | 1.2 (1.1, 1.3) | 1.1 (1.0, 1.2) |
| No | 50.3 | 1.0 | 1.0 | 46.2 | 1.0 | 1.0 |
| **Reduced household ability to buy food** |  |  |  |  |  |  |
| A lot | 66.9 | 1.6 (1.4, 1.9) | 1.4 (1.2, 1.6) | 61.1 | 1.9 (1.6, 2.2) | 1.6 (1.3, 1.9) |
| Somewhat | 54.8 | 1.3 (1.2, 1.6) | 1.2 (1.0, 1.4) | 52.5 | 1.6 (1.4, 1.9) | 1.5 (1.2, 1.7) |
| A little | 50.8 | 1.2 (1.1, 1.4) | 1.2 (1.0, 1.4) | 42.2 | 1.3 (1.1, 1.6) | 1.2 (1.0, 1.5) |
| Not at all/made it easier | 41.1 | 1.0 | 1.0 | 32.6 | 1.0 | 1.0 |
| **How worried about losing housing** |  |  |  |  |  |  |
| Very worried | 72.2 | 1.5 (1.4, 1.7) | 1.3 (1.2, 1.4) | 62.5 | 1.5 (1.4, 1.7) | 1.3 (1.1, 1.4) |
| Somewhat worried | 58.3 | 1.2 (1.1, 1.4) | 1.1 (1.0, 1.3) | 55.2 | 1.3 (1.2, 1.5) | 1.2 (1.1, 1.4) |
| Not worried at all | 47.2 | 1.0 | 1.0 | 41.1 | 1.0 | 1.0 |
| **Expect to graduate** |  |  |  |  |  |  |
| Later than expected | 66.7 | 1.4 (1.3, 1.6) | 1.3 (1.1, 1.4) | 59.3 | 1.3 (1.2, 1.5) | 1.1 (1.0, 1.3) |
| Don't know | 55.6 | 1.2 (1.1, 1.3) | 1.1 (1.0, 1.2) | 46.6 | 1.0 (0.9, 1.2) | 1.0 (0.9, 1.1) |
| Earlier/same time as expected | 46.7 | 1.0 | 1.0 | 44.5 | 1.0 | 1.0 |

PR=prevalence ratio, CI=confidence interval

The synergistic impact of financial stressors, health of family members, and one’s own health on respondents’ mental health was also evident in many of the open-ended responses. One student explained the relationship between mental health and financial stressors as follows, “Being fired from my job gives me plenty of free time but causes many panic attacks. I am very stressed and scared for my health and my family as well. Financially we’ve been struggling a lot.” In talking about the health of family members, a student explained, “I find it extremely difficult to concentrate because I am in constant fear my child or my elderly parents are going to die…” Another student described coping with loss, “My father passed away because of the virus. It has been really hard because he was my everything…” One student described why the virus was stressful for her, “During the time I believed I had the virus, I was very worried and unable to focus. Now is mostly a concern about how long this will go on and will it affect my ability to take care of my family financially.”

## Discussion

In April 2020, the prevalence of anxiety and depression was high among CUNY students—more than double the estimate of these conditions from a representative survey of CUNY students in 2018.^13^ This finding is consistent with national studies indicating elevated levels of depression and anxiety among adults as a result of the COVID-19 pandemic.^14^ Furthermore, CUNY students reported high levels of economic instability, with a high percentage reporting loss of household income, concerns about housing, and food insecurity. Many students were also concerned about delays to graduation as a result of the pandemic. Previous studies have shown that food insecurity, depression and anxiety can undermine academic success.^15-17^ The significant increase in these conditions among college students found in our survey and other studies on the impact of the COVID-19^18^ suggest that the pandemic may disrupt the progress of many students in completing their college degrees, a credential that offers lifetime health, economic and social benefits.^19^

In multivariable models, concerns about the ability to pay for housing and food, the student or someone in their household experiencing possible COVID-19 symptoms, and caretaking for adults over the age of 65 were associated with worse mental health outcomes. These findings are consistent with a study of medical college students in China during the COVID-19 pandemic, which found that family economic stability was associated with reduced levels of anxiety, and being sick or having a family member be sick were associated with increased anxiety levels.^20^ Findings are also consistent with a study conducted by the US Centers for Disease Control and Prevention (CDC), which found that, for young adults, being an unpaid caretaker was associated with poor mental health outcomes during the first wave of the COVID-19 pandemic.^14^ Females generally have higher rates of anxiety/depression then males in the general population^21^ and tend to have higher caretaking responsibilities^22^ which may explain our finding that females were at higher risk for anxiety/depression than males.

Our study has a number of limitations. Recall bias may have affected students’ reports on general health at the beginning of the semester. We collected cross-sectional data in April 2020 prior to the first stimulus bill, which temporarily increased unemployment benefits. The financial impact of the COVID-19 pandemic is changing, and students may be experiencing different levels of financial instability given the temporal changes to available benefits. We had a 23% response rate, which may introduce bias in our results, even with the creation of non-response weights. Finally, our study is cross-sectional, and thus factors associated with mental health outcomes may not be causal. We note, however, that there is a plausible mechanism by which financial stressors, dealing with COVID-19 illness, being a caretaker and worries about delays in graduation could lead to poor mental health and many of the open-ended responses support the direction of these associations.

Our results suggest that to mitigate the negative impact of the COVID-19 pandemic on low-income urban college students requires a multi-pronged approach. First, universities need to ensure that students have access to campus, community and telehealth mental health services. In our study, students with anxiety/depression were more likely to report increased substance use and increased anxiety around delays to graduation, than those not experiencing anxiety/depression, highlighting the importance of reaching these students. Even before the pandemic, college students and other young adults often had limited access to affordable mental health services;^23^ this problem likely has been exacerbated by COVID-19. Despite the need to ensure remote services during COVID-19, a review of 138 college websites in the NYC metropolitan area found that only half included information on how students could access remote mental health services.^24^

Ensuring access to mental health services may not be sufficient to address high rates of anxiety/depression without also addressing basic needs. The fact that a sizable proportion of CUNY students are experiencing financial instability including fear of losing housing and food insecurity needs to be addressed. These findings likely apply to many college students in public universities in the United States. A study of 651 college students in a large state-funded university in Texas, found 34.5% of their students reported food insecurity in the month prior to the survey, with those indicating loss of income and changes in living situations due to the COVID-19 pandemic at the highest risk for food insecurity.^25^ Interventions to address these concerns include expansion of federal eligibility and benefit levels, simplifying the process for college students to enroll in Supplemental Nutrition Assistance Program (SNAP) and other public benefits, increasing access to financial aid, moving toward free college tuition, maintaining supplemental cost of living expenses to unemployment benefits and/or implementing a universal basic income.

Delays to graduation caused by the pandemic at institutions such as CUNY, which serves a high proportion of low income and Black and Hispanic students, could further exacerbate existing economic disparities and social inequities. Implementation of new policies and programs to increase college student access to mental health services and public benefits can help to prevent these inequities from growing. Finally, while becoming infected with COVID-19 takes a toll on students and their loved ones physically, the mental health aspect of becoming infected or thinking one may be infected cannot be ignored. We found a substantial increase in students reporting their general health was poor or fair as a result of the pandemic.

### Public Health Implications

Much of the public discussion of COVID-19 on campuses has emphasized the role of the pandemic on community transmission of infection.^26^ However, the majority of US college students do not attend residential campuses, but rather go to community colleges or local campuses and live at home with their families or peers.^27^ As a population, CUNY students more resemble the majority of US college students than those attending private institutions with on-campus housing. Thus, preventing further increases to and mitigating the consequences of spikes in food and housing insecurity, depression, and anxiety among college students such as those at CUNY can contribute both to mitigating the effects of the pandemic and to achieving national health and equity goals.

## Data Availability

Not yet publicly available.

